# Sensitivity of Nasopharyngeal, Nasal and Throat Swab for the Detection of SARS-CoV-2

**DOI:** 10.1101/2020.05.05.20084889

**Authors:** Byron M. Berenger, Kevin Fonseca, Angela R. Schneider, Jia Hu, Nathan Zelyas

## Abstract

Nasopharyngeal (NP), nasal and throat swabs are the most practical specimen sources to test for upper respiratory pathogens. We compared the sensitivity of NP, nasal and throat swabs to detect SARS-CoV-2 in community patients. Using detection at any site as the standard, the sensitivities were 90%, 80% and 87% for NP, nasal and throat respectively (n=30 positive at any site). Throat swabs are likely a suitable alternative to NP swabs for the detection of COVID-19 infections.

## Introduction

The specimen source of choice for the screening of coronavirus disease-2019 (COVID-19) varies globally. Generally, the ideal specimen type for viruses causing respiratory tract infections is a nasopharyngeal (NP) swab (1). Lower respiratory tract specimens may be of benefit in severe cases of COVID-19, but most cases have mild upper respiratory tract disease (2–4). However, due to worldwide shortages of swabs and collection media, it has become necessary to identify alternate methods to NP swabs for sample collection for COVID-19 testing. We enrolled COVID-19 positive community patients on home isolation to determine the sensitivity of NP, nasal and throat swabs.

## Methods

Alberta Health Services (AHS) Public Health provided a list of people who had tested positive for COVID-19. Oral consent by phone was obtained for collection of NP, nasal, and throat swabs in the participant’s home. NP swabs were collected using the Flexible Mini Tip Flocked Swab (Copan S.P.A, Italy) in Universal Transport Media (UTM, Copan), nasal swabs using APTIMA Unisex Collection Kit (Hologic Inc., Marlborough, Mass), and throat swabs using the APTIMA Multitest Collection Kit. Collectors were given instructions on how to perform swabs. For NP swabs the AHS collection guide was used (5). For nasal collection, both nares were swabbed to a depth of at least 3 cm (or until resistance felt) and rotated three times. Throat swabs were collected from both sides of the oropharynx and the posterior pharyngeal wall under the uvula. The University of Calgary Research Ethics board approved this study (REB20-444).

Testing for SARS-CoV-2 was performed by a multiplex reverse transcriptase real time-polymerase chain reaction (RT-PCR). The RT-PCR was developed, validated and performed at the Alberta Public Health Laboratory (ProvLab) targeting the envelope region (modified from (6)) and the RNA-dependent RNA polymerase encoding regions (E and RdRp genes, respectively). The test was validated against proficiency panels provided by the Public Health Agency of Canada National Microbiology Laboratory (Winnipeg, MB). Spiking APTIMA media with a positive NP swab UTM specimen showed minimal difference in the Ct values for our SARS-CoV-2 PCR and a 2 log dilution series in APTIMA, stored at 4°C and room temperature for 48 h, showed minimal difference from time zero (≤0.6 change in Ct value). Graph Pad Prism v8.4.1 (Graphpad Prism Softwar L.L.C, San Diego, CA) was used for statistical analysis.

## Results

Of 82 COVID-19 positive individuals contacted, 36 consented (41% female, mean age 44.6 (range 18-61)). Initial diagnosis was made by NP (n=15) or nasal swab (n=21). Participants were swabbed for the study a mean of 4.1 days (range 1 to 6) after the initial diagnostic test and 10 days (range 4 to 23) after symptom onset. Thirty of the thirty-six participants tested positive again at one or more of the three sites swabbed. The mean time from symptom onset and study swabs was 12.6 days (range 5-18) for those testing negative at all three sites and compared to 10.0 days (range 4-23) for those with a positive result at any site. Using a reference standard of a positive result at any site, NP swab had a sensitivity of 90% (95%CI 74.4-96.5), throat swab 87% (70.3-94.7) and nasal swab 80% (62.7-90.5) (Wilson/Brown Method, Table 1). In only two cases was only one specimen positive (both nasal). Seven participants were positive from only two sources (n=2 NP and nasal, n=5 NP and throat).

**Table 1a.**
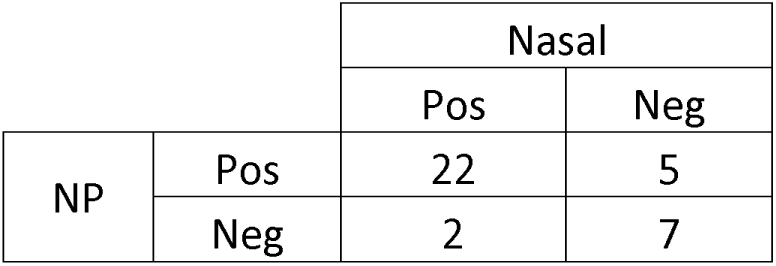
COVID-19 PCR Results for NP and Nasal swabs

|  |  | Nasal |  |
| --- | --- | --- | --- |
|  |  | Pos | Neg |
| NP | Pos | 22 | 5 |
|  | Neg | 2 | 7 |

**Table 1b.** COVID-19 PCR Results for Nasal and Throat Swabs

|  |  | Throat |  |
| --- | --- | --- | --- |
|  |  | Pos | Neg |
| Nasal | Pos | 20 | 4 |
|  | Neg | 6 | 6 |

**Table 1c.** COVID-19 PCR Results for NP and Throat Swabs

|  |  | Throat |  |
| --- | --- | --- | --- |
|  |  | Pos | Neg |
| NP | Pos | 25 | 2 |
|  | Neg | 1 | 8 |

Comparing the samples where all targets were positive and Ct values were available (n=19 for E gene and n=18 RdRp), the Ct values for NP swabs was lower than throat swabs for the E gene (p=0.028, p>0.22 for other site comparisons) (Friedman Test). The median Ct values for the E gene were 25.5 (10^th^ to −90^th^ percentile: 20.5-29.5) for NP, 27.6 (24.7-32.4) for nasal and 28.7 (23.5-34.2) for throat; median Ct values for the RdRp gene were 27.9 (23.5-32.4), 30.5 (27.5-35.0), and 31.3 (26.5-35.5), for the same sites, respectively (p>0.09).

## Discussion

Our study demonstrates that the sensitivity of nasal swabs is somewhat inferior to NP or throat swabs whereas throat and NP swabs have comparable sensitivity. This finding was despite the Ct value being higher in throat swabs compared to NP swabs. Consequently, when NP swabs are not available, throat swabs are a preferable alternative to nasal swabs for COVID-19 testing.

Péré *et al* also found nasal swabs to be less sensitive than NP swabs (8). They reported a sensitivity of 89.2% (4/37 NP positives were false negative) with NP swabs as the reference standard. Using NP swabs as the reference standard in our study, the sensitivity of nasal swabs was 82.5% (5/27 NP positives were false negative). It is important to note that in our study NP swabs missed 3 positives that other sources detected (n=2 detected by nasal and n=1 throat). Combining results from Péré *et al* and our study gives a sensitivity for nasal swabs of 85.9%. Differences between studies included a different patient population (patients seen in hospital vs. community), PCR assay used and collection media. Although our study represents the general population with COVID-19, the results may differ in inpatients as they may have higher viral loads (9).

Contrary to our findings, Wang *et al* (10) reported 73% of patients with a positive NP swab result tested negative by throat/oropharyngeal swab. A potential explanation is that Wang *et al* did not instruct collectors to swab under the epiglottis. Therefore, viral shedding from the nasopharynx may have not been optimally sampled in the Wang *et al* cohort.

One limitation of our study is the small number of samples validated. We chose thirty to have a high likelihood to detect 90% agreement (11). A hindrance of performing studies of this nature with a large sample size is that sampling puts collectors at risk of infection even with appropriate personal protective equipment. Our study was also performed a mean of 10 days after symptom onset, so the site of optimal sampling may have differed if participants were swabbed closer to symptom onset. In early disease, throat swabs may be falsely negative compared to CT scan findings, though it is not clear whether NP and nasal swabs also lack sensitivity early in the disease progress (12).

The sensitivity of the sample type is dependent on proper sampling procedure. In our jurisdiction, nasal swabs were initially implemented due to reports of lower Ct values than those seen in throat swabs (13). Despite education and routine observation of the technique used at COVID-19 community assessment centres, multiple accounts of sampling the anterior nares instead of the posterior nares/lower turbinates were reported to the laboratory by patients. Sampling errors may also occur with throat and NP swab and may have contributed to some of the false-negative NP swab results despite our collectors being trained health care professionals. Additionally, other studies have since reported that throat swabs have equivalent or higher SARS-CoV-2 viral loads compared to NP or nasal swabs, respectively (14, 15). Based on our results and familiarity of health care providers with throat swabs (as opposed to nasal swabs), we currently recommend in our jurisdiction the collection of throat swabs if NP swabs are not available.

## Data Availability

Data is available on request by contacting the corresponding author.

## Acknowledgements

This study was performed using operational funds of Alberta Health Services (AHS) and Alberta Precision Laboratories. AHS Mobile Integrated Health Care for collecting the samples, particularly Ryan Kozicky and Michel Smith for coordinating. ProvLab for supporting the study and the ProvLab staff for running the tests.

